# Non-glycemic genetic effects on HbA1c and clinical glycemic status in African ancestry: VA Million Veteran Program

**DOI:** 10.1101/2024.05.26.24307947

**Authors:** Laurel Stell, Kent Heberer, Kyung Min Lee, Shoa Clarke, Wu Fan, Ilana Belitskaya-Levy, Julie A. Lynch, Hua Tang, Marijana Vujkovic, Donald Miller, Themistocles L. Assimes, Phil Tsao, Scott Damrauer, Kyong-mi Chang, Jennifer S. Lee, the VA Million Veteran Program

**Author notes:** Corresponding author: Laurel Stell, Department of Biomedical Data Science, Stanford University School of Medicine, MSOB, 1265 Welch Rd, Room X3C67, Stanford, CA 94305. These authors contributed equally (Kent Heberer, Kyung Min Lee).

## Abstract

**IMPORTANCE:** Clinical guidelines recommend against using glycated hemoglobin A1c (HbA1c) to assess glycemia in patients with two erythropoietic conditions: glucose-6-phosphate dehydrogenase (G6PD) deficiency or sickle cell disease. What remains elusive is quantifying the impact of genetic variants underlying these and other erythropoietic conditions on HbA1c levels as a clinical indicator of glycemic status.

**OBJECTIVE:** To evaluate the impact of five erythropoietic (non-glycemic) genetic variants on HbA1c levels as a clinical indicator of glycemic status in a population with African genetic ancestry.

**DESIGN:** Retrospective cohort study conducted January 2011 to November 2022.

**SETTING:** Veterans Health Administration’s (VHA’s) Million Veteran Program (MVP), a genetic biobank representative of the U.S.’s largest integrated healthcare system, including the longest-running electronic health record system.

**PARTICIPANTS:** 84,987 MVP participants with African genetic ancestry, excluding those with type 1 or secondary diabetes or G6PD, sickle cell, or other erythropoietic conditions.

**EXPOSURE:** Any one of five erythropoietic genetic variants known or suspected to affect HbA1c levels in African ancestry.

**MAIN OUTCOMES AND MEASURES:** Clinically measured HbA1c, random blood glucose, triglyceride to HDL-cholesterol ratio, body mass index, blood pressure, diagnosis of and medications for type 2 diabetes (T2D).

**RESULTS:** All the variants had significant differences in HbA1c compared to non-carriers with the same sex, age, glucose and BMI. Males with X-linked G202A variant for G6PD deficiency (11% of males in cohort) had HbA1c levels reduced by 0.8 percentage points compared to non-carriers; female carriers had reductions over 0.3 percentage points. Overall, G202A carriers had worse dysglycemia but were less likely to be diagnosed with or prescribed medication for dysglycemia than non-carriers. Due to this variant, an estimated 1.5 million (6%) African American adults without T2D may have dysglycemia in the pre-diabetes or T2D range, but their HbA1c indicate normoglycemia.

**CONCLUSIONS AND RELEVANCE:** By lowering HbA1c without lowering glucose level, G202A variant for G6PD deficiency—a condition common to African ancestry and largely asymptomatic and undiagnosed—could be misguiding clinical management for 9% of African American Veterans without T2D and 6% of African American adults without T2D in the U.S. and contributing to racial disparities in T2D management.

**KEY POINTS:** *QUESTION:* What is the impact of non-glycemic genetic variants on HbA1c levels as a clinical indicator of glycemic status?

*FINDINGS:* In patients with African ancestry within a U.S. healthcare setting, carriers of G202A variant for glucose-6-phosphate dehydrogenase (G6PD) deficiency have significantly lower HbA1c—0.9% points in males—and lower rates of type 2 diabetes diagnosis and medication but higher glucose compared to non-carriers. Four additional erythropoietic variants also had significant effects on HbA1c but none on glucose.

*MEANING:* HbA1c can underestimate dysglycemia in carriers of G202A variant, which is common in African, and very rare in European, ancestry.

## INTRODUCTION

Circulating glycated hemoglobin (HbA1c) is widely used to screen, diagnose, and monitor glycemic status: normoglycemia, pre-diabetes, and diabetes. It reflects glucose bound to hemoglobin over the life span of the erythrocyte (about 120 days). It consequently has less daily fluctuation and offers greater testing convenience than other clinical indicators of glycemic status such as blood glucose in the fasting state or after an oral glucose load. Clinical outcomes have been shown to be more strongly associated with HbA1c than with other standard glycemic measures.^1^ Yet unlike measures that rely on blood glucose, HbA1c can be affected by factors that disrupt function or metabolism of red blood cells.^2–5^

According to the American Diabetes Association (ADA) Standards of Medical Care (SOC) in Diabetes, HbA1c is an inaccurate indicator of glycemic status for patients with either of two erythropoietic conditions: G6PD deficiency or sickle cell disease.^5^ The chromosome X-linked G202A variant, which contributes to G6PD (A-) deficiency, has been reported to have relatively large non-glycemic effects on HbA1c levels.^4,6–9^ Other erythropoietic genetic variants also have been reported to have large effects on HbA1c: a secondary variant in *G6PD*,^8^ hemoglobin variant HbS which causes sickle cell disease,^7,9–11^ and a variant associated with alpha thalassemia (-α3.7 kb deletion).^9,11,12^ The hemoglobin variant HbC also has been suspected of affecting HbA1c levels.^11,13^

It is unclear whether G202A or these other variants impact clinical management of glycemic status. Most are common among individuals with African genetic ancestry; all are rare in individuals with European ancestry. The ADA SOC notes, “There is controversy regarding racial differences in A1C… There is an emerging understanding of the genetic determinants of A1C, but the field lacks adequate genetic data in diverse populations.”^4,14,15^

We analyzed the effect of these five variants using clinical and genetic data from 84,987 Million Veteran Program (MVP) participants with African genetic ancestry in the Veterans Health Administration (VHA). The VHA is the U.S.’s largest integrated healthcare system and has the longest-running electronic health record (EHR) system. Comparing individuals with one minor allele of any one of the variants to individuals carrying none (non-carriers), we analyzed the effects of the variants on clinically measured HbA1c levels, blood glucose levels, rates of T2D diagnosis and medication prescribing, and clinical indicators of insulin resistance at given HbA1c levels. In the VHA patient population and U.S. overall, we estimated the number of African American adults without T2D whose glycemic status determined by HbA1c might be misguided due to the presence of G202A variant.

## METHODS

### Study population and data

The study population is from MVP, described elsewhere.^16^ Briefly, U.S. veterans were recruited from 63 participating VHA medical facilities. At enrollment, which started in 2011, study participants provided blood samples for DNA extraction and genotyping and consent to access their VHA EHR for research purposes. The MVP and this study received ethical and study protocol approval from the VA Central Institutional Board (cIRB). MVP has conducted genotyping, imputation (using 1000 Genomes and African Genome Resources reference panels), and classification by genetically inferred ancestry for over 650,000 participants.^17,18^

The cohort for this study included individuals with African genetic ancestry. Because we extrapolated our results to other populations identified by self-identified race and without genetic data, we also required self-identified race “Black or African American”, which was 94.0% of African genetic ancestry. HbA1c, random blood glucose, HDL-cholesterol and triglyceride values were obtained from VHA’s Corporate Data Warehouse (CDW)^19^ of EHR (eTable 1 in Supplement 1). For each individual, the index date was the first date on or after MVP enrollment with values for all four of these labs. We excluded individuals (a) who did not have an index date, (b) with diagnostic codes on or before index date for type 1 diabetes or secondary causes of diabetes or for erythropoietic disorders such as sickle cell trait or G6PD deficiency (eTable 2 in Supplement 1) or (c) whose genetic sex differed from their self-identified gender, resulting in a cohort of 84,987 (72,604 male; eFigure 1 in Supplement 2).

When available, we obtained the following from CDW: height, weight within ±365 days of index date, two blood pressure readings on different days and within ±365 days of index date, ICD-9 or -10 diagnosis code for type 2 diabetes (T2D) on or before the index date, and medications for diabetes, lipid conditions, or hypertension on or within 365 days before index date (eTable 3 in Supplement 1). All clinical data were in outpatient settings, except diagnoses of erythropoietic disorders could be inpatient or outpatient. We categorized insulin resistance features—blood glucose, triglyceride to HDL-cholesterol ratio (TG/HDLC), body mass index (BMI) and blood pressure—as normal, elevated, high, or very high using published ranges (eTable 4 in Supplement 1; eMethods in Supplement 2). The level for blood pressure was obtained from the lower of the two readings. Individuals with TG/HDLC in normal range but with a medication for a lipid condition within one year before index date were in a separate category; an analogous approach was taken for individuals with normal blood pressure but with hypertensive medication.

Estimation of the effects of the five genetic variants on HbA1c was based on 76,865 individuals from this cohort in six non-overlapping groups: individuals with none of the variants (non-carriers) and individuals with exactly one minor allele among the variants (Table 1; eTable 5 in Supplement 1).

**Table 1.**
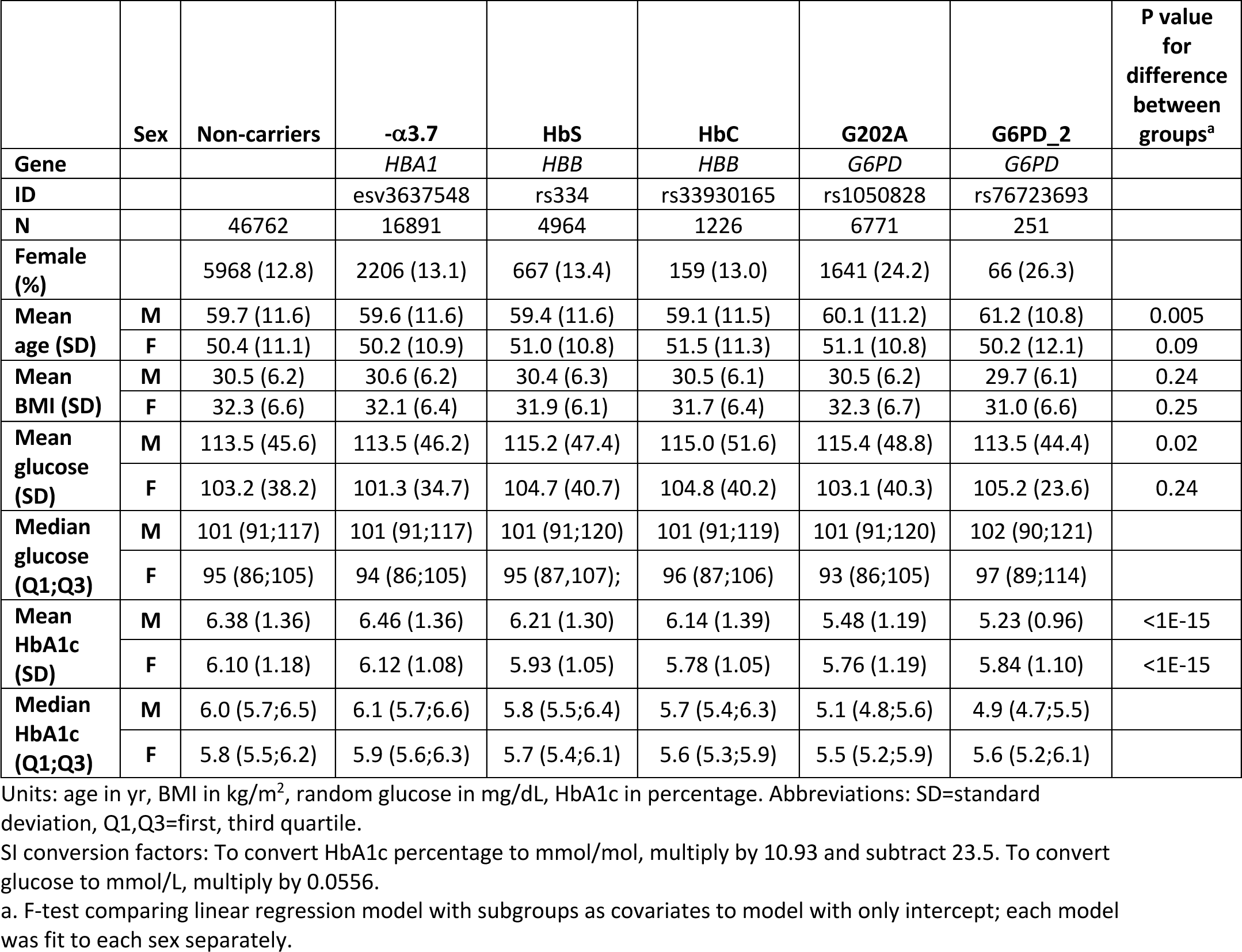
Characteristics of genetic variant groups.

| | Sex | Non-carriers | -α3.7 | HbS | HbC | G202A | G6PD_2 | P value for difference between groups<sup>a</sup> |
| --- | --- | --- | --- | --- | --- | --- | --- | --- |
| Gene |  |  | *HBA1* | *HBB* | *HBB* | *G6PD* | *G6PD* |  |
| ID |  |  | esv3637548 | rs334 | rs33930165 | rs1050828 | rs76723693 |  |
| N |  | 46762 | 16891 | 4964 | 1226 | 6771 | 251 |  |
| Female (%) |  | 5968 (12.8) | 2206 (13.1) | 667 (13.4) | 159 (13.0) | 1641 (24.2) | 66 (26.3) |  |
| Mean age (SD) | M | 59.7 (11.6) | 59.6 (11.6) | 59.4 (11.6) | 59.1 (11.5) | 60.1 (11.2) | 61.2 (10.8) | 0.005 |
|  | F | 50.4 (11.1) | 50.2 (10.9) | 51.0 (10.8) | 51.5 (11.3) | 51.1 (10.8) | 50.2 (12.1) | 0.09 |
| Mean BMI (SD) | M | 30.5 (6.2) | 30.6 (6.2) | 30.4 (6.3) | 30.5 (6.1) | 30.5 (6.2) | 29.7 (6.1) | 0.24 |
|  | F | 32.3 (6.6) | 32.1 (6.4) | 31.9 (6.1) | 31.7 (6.4) | 32.3 (6.7) | 31.0 (6.6) | 0.25 |
| Mean glucose (SD) | M | 113.5 (45.6) | 113.5 (46.2) | 115.2 (47.4) | 115.0 (51.6) | 115.4 (48.8) | 113.5 (44.4) | 0.02 |
|  | F | 103.2 (38.2) | 101.3 (34.7) | 104.7 (40.7) | 104.8 (40.2) | 103.1 (40.3) | 105.2 (23.6) | 0.24 |
| Median glucose (Q1;Q3) | M | 101 (91;117) | 101 (91;117) | 101 (91;120) | 101 (91;119) | 101 (91;120) | 102 (90;121) |  |
|  | F | 95 (86;105) | 94 (86;105) | 95 (87;107); | 96 (87;106) | 93 (86;105) | 97 (89;114) |  |
| Mean HbA1c (SD) | M | 6.38 (1.36) | 6.46 (1.36) | 6.21 (1.30) | 6.14 (1.39) | 5.48 (1.19) | 5.23 (0.96) | <1E-15 |
|  | F | 6.10 (1.18) | 6.12 (1.08) | 5.93 (1.05) | 5.78 (1.05) | 5.76 (1.19) | 5.84 (1.10) | <1E-15 |
| Median HbA1c (Q1;Q3) | M | 6.0 (5.7;6.5) | 6.1 (5.7;6.6) | 5.8 (5.5;6.4) | 5.7 (5.4;6.3) | 5.1 (4.8;5.6) | 4.9 (4.7;5.5) |  |
|  | F | 5.8 (5.5;6.2) | 5.9 (5.6;6.3) | 5.7 (5.4;6.1) | 5.6 (5.3;5.9) | 5.5 (5.2;5.9) | 5.6 (5.2;6.1) |  |

Units: age in yr, BMI in kg/m<sup>2</sup>, random glucose in mg/dL, HbA1c in percentage. Abbreviations: SD=standard deviation, Q1,Q3=first, third quartile.
SI conversion factors: To convert HbA1c percentage to mmol/mol, multiply by 10.93 and subtract 23.5. To convert glucose to mmol/L, multiply by 0.0556.
a. F-test comparing linear regression model with subgroups as covariates to model with only intercept; each model was fit to each sex separately.

### Statistical analysis

All analyses were performed with R software (version 4.0; R Foundation for Statistical Computing, Vienna, Austria). Estimates of genetic effects and their P values are from regression with covariates including age, sex, variant group and variant-sex cross terms (referred to as the “basic covariates”). We estimated (a) the effect in male carriers compared to male non-carriers and (b) the change in effect size between sexes. For each fit, we required P<0.01 for statistical significance to account for testing five variants.

To test genetic effects on the relationship between HbA1c and glucose, we performed linear regression of HbA1c with covariates glucose, glucose squared and BMI in addition to the basic covariates, excluding individuals with T2D diagnosis or medication. To estimate the odds ratio (OR) comparing carriers to non-carriers for binary outcomes, we used logistic regression with the basic covariates and, for outcomes T2D diagnosis and medication, glucose and BMI. To test whether HbA1c or glucose was a better indicator of insulin resistance features, we performed linear regression of each feature with with the basic covariates and either (a) HbA1c and its square or (b) glucose and its square, excluding individuals with medications for T2D, lipid conditions or hypertension. Significance of a variant indicates need for a genetic adjustment in order for HbA1c or glucose to predict the feature.

Based on our results, we defined mis-designations of glycemic status—normoglycemia, pre-diabetes or T2D—based on HbA1c. To estimate mis-designations in the current VHA patient population (as of June 7, 2023), we analyzed the most recent HbA1c value not before 2018 from curated VHA EHR data in the CDW for individuals identified as “Black or African American” in at least one record, excluding patients with known diabetes diagnosis. To estimate mis-designations in the U.S. overall, we analyzed data for “non-Hispanic Black” adults with neither previous diabetes diagnosis nor current diabetes medication from the 2017–2018 National Health and Nutrition Examination Survey (NHANES), a nationally representative sample of U.S. residents, following NHANES analysis guidelines including use of survey package in R and categorizing age into three levels: younger than 40 years, 40-59 and 60 or older.^20^ We used the observed HbA1c distribution in the VHA population and the estimation from NHANES for the U.S. To estimate the proportion that would have the G202A variant at given HbA1c level, we used the same proportion as in our MVP cohort after excluding T2D diagnosis or medication (eMethods and eFigure 1 in Supplement 2).

## RESULTS

In the variant groups including 76,865 individuals, males comprised 86% of the study cohort, but the cohort included more than 10,000 females (characteristics of the groups in Table 1). G202A in *G6PD* gene and -α3.7 in *HBA1* were common erythropoietic variants (minor allele frequency (MAF) 11-15%), followed by HbS and HbC in *HBB* gene (MAF 1-5%) and the rare *G6PD* secondary variant (MAF 0.4%). These MAFs were slightly lower than in the overall MVP cohort with African ancestry (eTable 5 in Supplement 1).

The two X-linked *G6PD* variants had the largest magnitude of effect on HbA1c level (Figure 1A). Specifically, adjusting for age, glucose and BMI, carriers of the more frequent of the two *G6PD* variants, G202A, had HbA1c 0.8-percentage points lower than non-carriers in males and 0.3-percentage points lower than non-carriers in females (Table 2A). Although G202A’s magnitude of effect in females was half that in males, it was still larger than any of the autosomal variants.

**Figure 1.**
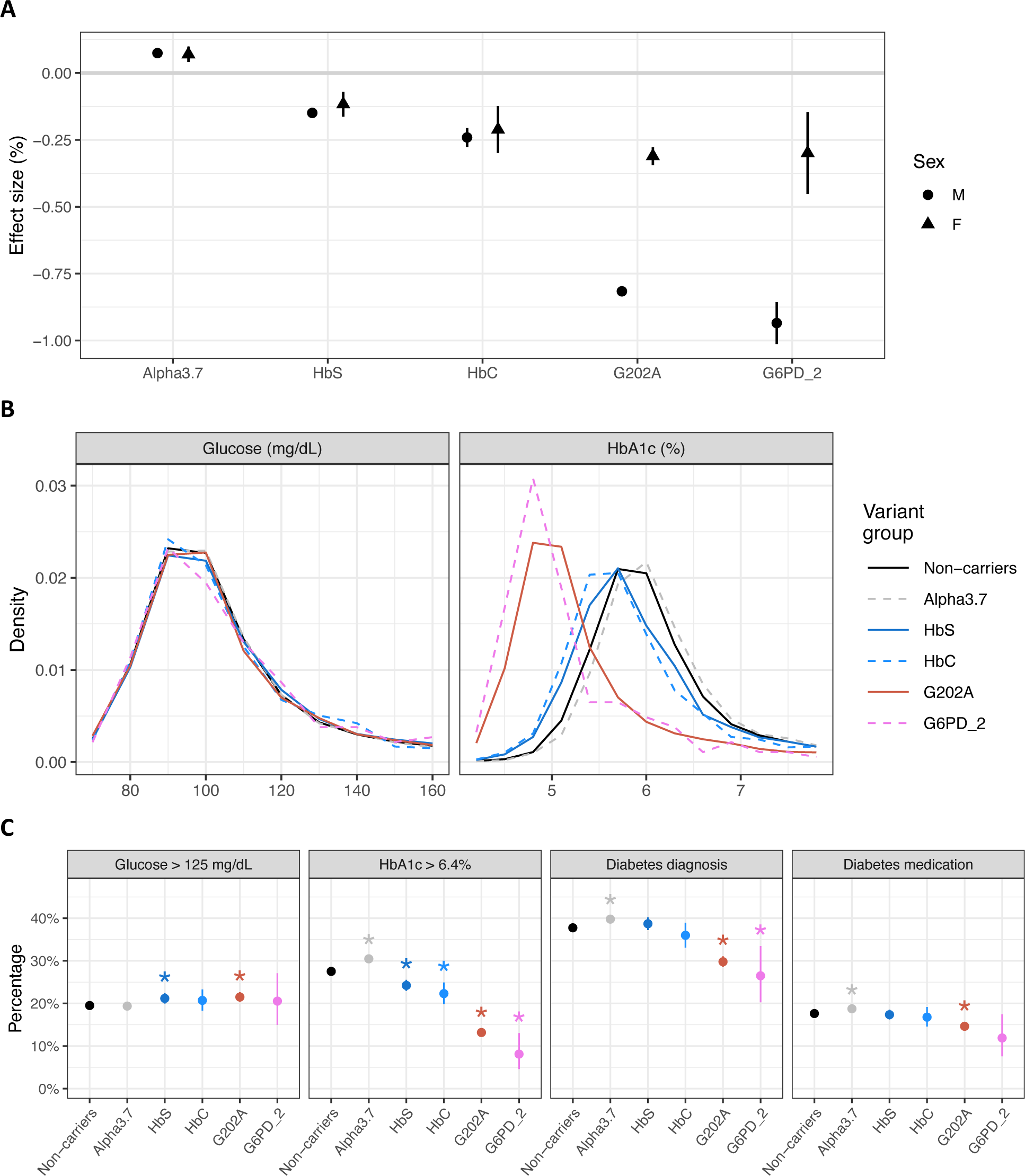
Differences between carriers and non-carriers in HbA1c, random blood glucose, T2D diagnosis and T2D medication. (A) Genetic effects on HbA1c after adjusting for age, sex, glucose and BMI, with vertical lines indicating ±2 standard errors. (B) Distributions of random glucose and HbA1c in males, grouped by variant. (Distributions by age group and sex are in eFigures 2,3 in Supplement 2.) (C) Percentage of each variant group satisfying selected T2D-related features in males, with vertical lines indicating 95% confidence intervals and an asterisk above a bar indicating P<0.01 for odds different in carriers compared to non-carriers (Table 3).

**Table 2.** Genetic effects on HbA1c and random blood glucose from linear regression. For females, rows omitted when variant-sex cross term not significant (P>0.01); value in “Difference” column is coefficient of variant-sex cross term added to effect size in males.

| **A. Linear regression of HbA1c with basic covariates plus glucose, glucose squared and BMI, excluding for known T2D diagnosis or medication** |  |  |  |
| --- | --- | --- | --- |
| **Variant** | **Difference, %** | **SE, %** | **P value** |
| **Males** |  |  |  |
| -$\alpha 3.7$ | 0.074 | 0.006 | <1E-15 |
| HbS | -0.149 | 0.009 | <1E-15 |
| HbC | -0.241 | 0.018 | <1E-15 |
| G202A | -0.816 | 0.008 | <1E-15 |
| G6PD_2 | -0.935 | 0.039 | <1E-15 |
| **Females** (SE and P value from test of difference in variant effect size between sexes) |  |  |  |
| G202A | -0.311 | 0.017 | <1E-15 |
| G6PD_2 | -0.299 | 0.076 | <1E-15 |
| **B. HbA1c difference between carriers and non-carriers from linear regression with basic covariates** |  |  |  |
| **Variant** | **Difference, %** | **SE, %** | **P value** |
| **Males** |  |  |  |
| -$\alpha 3.7$ | 0.083 | 0.013 | 5E-11 |
| HbS | -0.165 | 0.021 | 3E-15 |
| HbC | -0.226 | 0.041 | 3E-08 |
| G202A | -0.903 | 0.019 | <1E-15 |
| G6PD_2 | -1.167 | 0.097 | <1E-15 |
| **Females** (SE and P value from test of difference in variant effect size between sexes) |  |  |  |
| G202A | -0.339 | 0.041 | <1E-15 |
| G6PD_2 | -0.257 | 0.189 | 1E-06 |
| **C. Glucose difference between carriers and non-carriers from linear regression with basic covariates** |  |  |  |
| **Variant** | **Difference, mg/dL** | **SE, mg/dL** | **P value** |
| **Males** |  |  |  |
| -$\alpha 3.7$ | 0.02 | 0.43 | 0.96 |
| HbS | 1.83 | 0.72 | 0.01 |
| HbC | 1.65 | 1.40 | 0.24 |
| G202A | 1.75 | 0.67 | 0.009 |
| G6PD_2 | -0.45 | 3.32 | 0.89 |

SE=standard error.
SI conversion factors: To convert HbA1c difference or SE in percentage to mmol/mol, multiply by 10.93. To convert glucose to mmol/L, multiply by 0.0556.

All variants were much more strongly associated with HbA1c than with random blood glucose (Table 2B,C; Figure 1B; eFigures 2,3 in Supplement 2). Carriers of each variant had different mean HbA1c compared to non-carriers of the same sex and age (P<1E-8). Carriers of -α3.7 had higher HbA1c while carriers of the other variants had lower HbA1c. Only HbS and G202A carriers had differences in mean random blood glucose (P≈0.01), and those carriers had higher glucose than non-carriers.

For all variants, carriers and non-carriers had different rates of HbA1c in the diabetes range (HbA1c ≥6.5%; Table 3A; Figure 1C). Notably, male G202A carriers had odds of HbA1c in the diabetes range 0.39 times lower than the odds for male non-carriers at the same age. On the other hand, male G202A carriers had a significantly higher rate of glucose >125 mg/dL (Table 3B; Figure 1C). Likewise, male HbS carriers had lower rates of HbA1c in diabetes range but higher rates of glucose >125 mg/dL. Male G202A carriers also had 0.72 times lower odds of being prescribed diabetes medication than non-carriers with the same age, glucose and BMI, and had 0.57 times lower odds of having a T2D ICD code (Table 3C,D; Figure 1C).

**Table 3.**
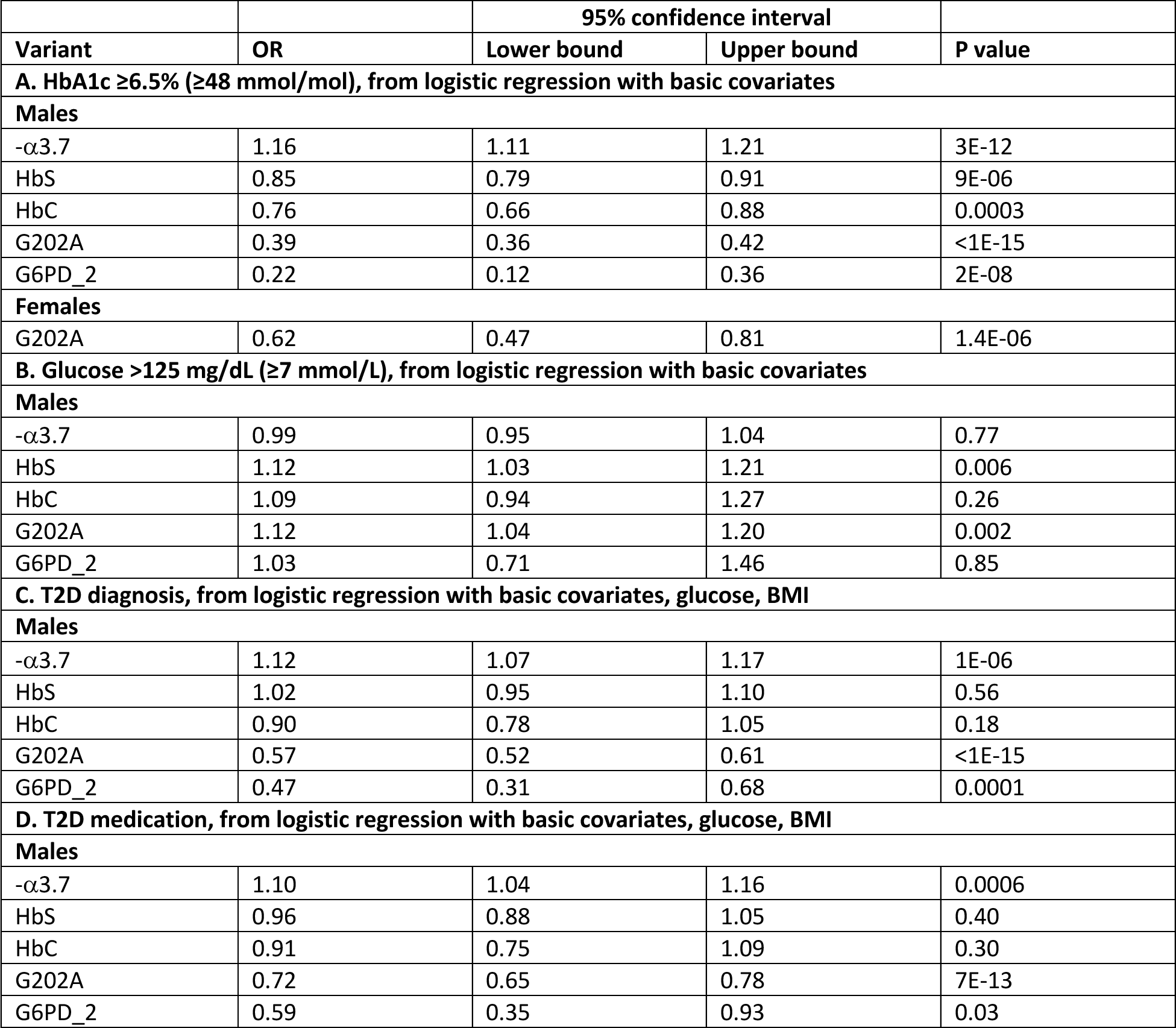
Odds ratios (ORs) comparing carriers to non-carriers from logistic regression of selected binary T2D-related outcomes. Results for females omitted because no variant-sex cross terms significant (P>0.01), except G202A and G6PD_2 in (A)—and G6PD_2 result omitted because its 95% confidence interval covers 1.

As with glucose >125 mg/dL, insulin resistance features were more frequent in G202A carriers than non-carriers. Male carriers of either *G6PD* variant who had HbA1c in pre-diabetes range (5.7-6.4%) had worse insulin resistance features than non-carriers in the same HbA1c range (Figure 2A; eFigure 4 in Supplement 2). Instead, the non-carriers with HbA1c in this range had insulin resistance features similar to G202A carriers with HbA1c 0.8-percentage points lower (4.9-5.6%; Figure 2B; eFigure 5 in Supplement 2). Likewise, female non-carriers with pre-diabetes HbA1c had insulin resistance features comparable to female G202A homozygotes with HbA1c 0.7-percentage points lower (eFigures 6,7 in Supplement 2). To quantify these visual relationships, we performed linear regression of each insulin resistance with covariates HbA1c and its square along with the basic covariates. G202A was significant in regression of TG/HDL, BMI or systolic blood pressure (P=8E-15, 6E-55, 4E-07, respectively; P=0.02 for diastolic blood pressure). With glucose in place of HbA1c, G202 was not significant for any of the four features (P>0.3). Hence, G202A affects the relationship of these insulin resistance features with HbA1c but not glucose.

**Figure 2.**
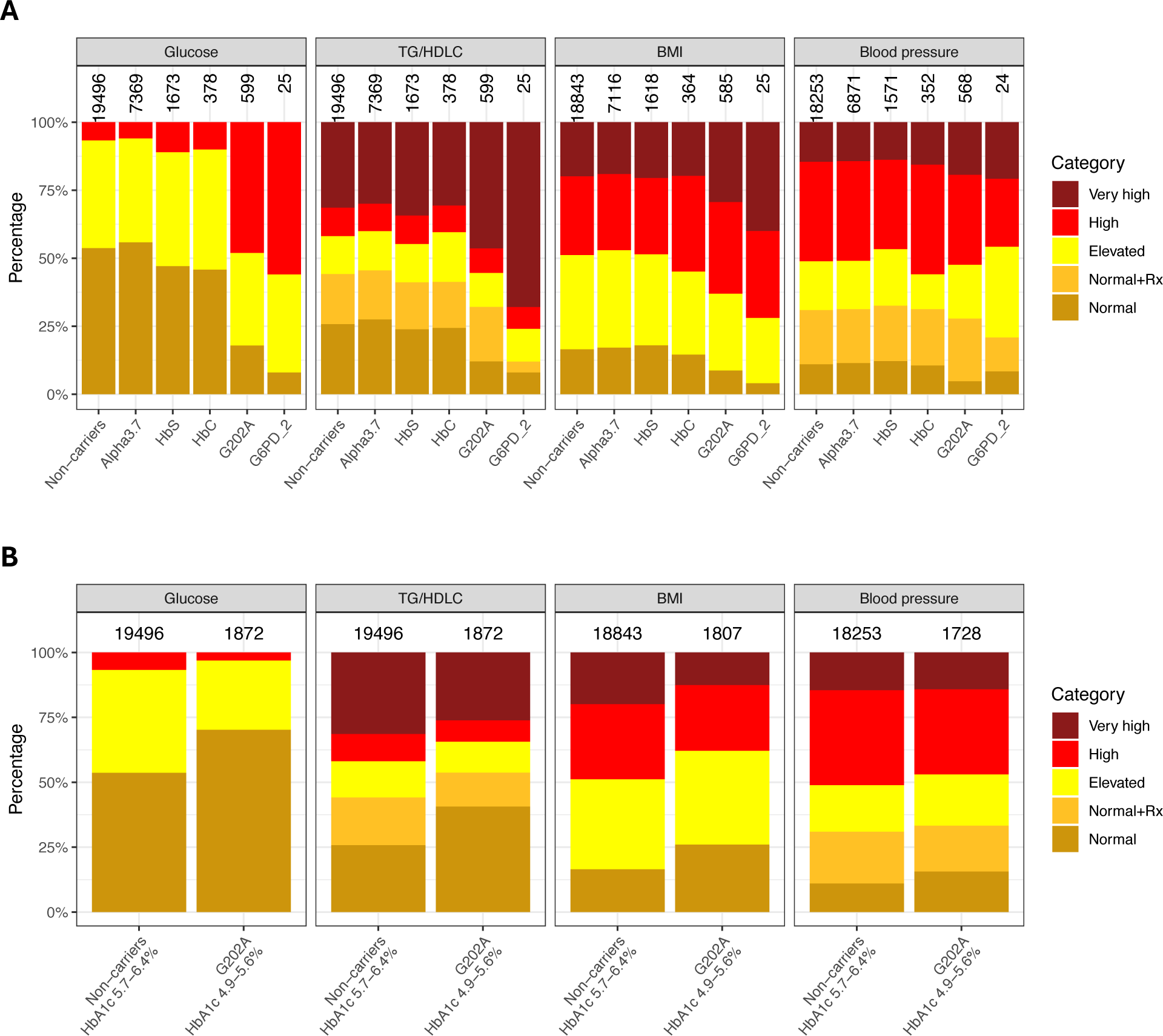
Comparison of insulin resistance features among males. (A) Distributions by variant group among individuals with HbA1c in pre-diabetes range (5.7-6.4%). (B) Distributions in non-carriers with HbA1c 5.7-6.4% compared to G202A carriers with HbA1c 4.9-5.6%. The number above each bar indicates the number of individuals represented. Categories of insulin resistance features defined in eTable 4 in Supplement 2.

We estimated the proportion of patients whose glycemic status might be misguided due to the G202A variant’s effect on HbA1c level (eTable 6A in Supplement 1). In 54,359 patients without known diabetes, an estimated 7.7% were in pre-diabetes but had an HbA1c in the normoglycemia range due to the G202A effect of lowering HbA1c (eTable 6B in Supplement 1). Another 1.0% were in T2D but had an HbA1c in the prediabetes range due to G202A. In the broader VHA patient population of 616,545 African American adults without diabetes, we estimated 51,000 (8.2%) were in pre-diabetes but had an HbA1c in the normoglycemia range due to G202A’s effect on HbA1c (eTable 6C in Supplement 1). Another 4,000 (0.7%) were in T2D but had an HbA1c in the pre-diabetes range due to G202A. Using NHANES to represent the overall U.S. population of African American adults without diabetes, we estimated 1.5 million (6.0%) to be in pre-diabetes but misguided by HbA1c as having normoglycemia and 125,000 (0.5%) to be in T2D but misguided by HbA1c level as having pre-diabetes, due to G202A’s effect on HbA1c (eTable 6D in Supplement 1).

## DISCUSSION

X-linked G202A, a common variant in African ancestry, had by far the greatest overall impact due to its combination of large effect size and common occurrence. Males with G202A had a 0.8-percentage points lower HbA1c, and females with one minor allele had a 0.3-percentage points lower HbA1c. Despite having higher rates of elevated glucose and insulin resistance features, such individuals had lower rates of T2D diagnosis and diabetes medication prescribing. This observation is consistent with G202A carriers being clinically managed as having better glycemic status than non-carriers. Rare *G6PD* variant rs76723693 in *G6PD* had effects on HbA1c even larger than G202A, while heterozygotes of -α3.7, HbS, or HbC variants had HbA1c altered by 0.2-percentage points or less. We observed these findings in the largest genotyped cohort of patients with African ancestry to date. Among African American adults without diabetes diagnosis, we estimated that 1.5 million (6.0%) are in pre-diabetes state but have a normoglycemic HbA1c result (<5.7%) due to the G202A variant.

Compared to Whites, African Americans have higher rates of T2D and some T2D-related complications, in addition to delays in interventions against glycemic dysregulation.^21,22^ Since it causes lower HbA1c results, G202A could be contributing to these disparities by underestimating severity of glycemic dysregulation in 11% of our cohort with African ancestry (after excluding for diagnosis of G6PD deficiency), while only <0.1% in Whites. Reassuringly, among African American men 40 years or older without diabetes, only less than 1% would be in T2D but misguided to be in pre-diabetes by HbA1c due to G202A.

To our knowledge, the significant effect of HbC on HbA1c levels has not been reported. Our effect estimates for the two *G6PD* variants and -α3.7 are consistent with prior estimates in U.S. African ancestry populations,^6,8,9^. Our HbS effect estimate on HbA1c of -0.14 percentage points since 2018 (eMethods in Supplement 2) was consistent with a recent report of -0.11 percentage points.^9^ We did not have information on HbA1c assay type, but our results support improvement in HbS assay interference even after HbA1c was recommended as a glycemic lab indicator of pre-diabetes and diabetes in 2010.^23^

Major strengths of this study are that the VHA is the U.S.’s largest national integrated healthcare system, has the longest running EHR system, and hosts the MVP, one of the largest multi-racial/ethnic genetic data sets in the world. Our cohort of 80,000 with African genetic ancestry dwarfs the sizes of previous studies, none of which exceeded 10,000. In contrast to non-healthcare system settings, other biobanks, or study-specific data sets, this study has clinical laboratory measurements including HbA1c, blood glucose, and lipid levels as well as electronic health data, including clinical diagnoses of diabetes and G6PD deficiency and T2D medication prescribing information.

Several potential study limitations should be considered. First, females were only 14% of the cohort. Even so, the study included 10,000 females, which is greater than the total number of both sexes in previous studies of African ancestry. Second, the VHA patient population skews older than the U.S. population, but we adjusted for age or used age stratification as appropriate in our analyses. Third, the blood glucose measurements in this study were not known to be fasting, which could result in overestimating glycemic dysregulation and insulin resistance; but our conclusions were reinforced by other clinical indicators of insulin resistance. Fourth, when estimating missed pre-diabetes or T2D due to G202A among African Americans without diabetes in the VHA population or in the U.S., we assumed that G202A frequency, as a function of HbA1c, sex and age, was the same as in our MVP cohort. Since MVP participants are largely representative of the VHA patient population (eFigure 9 in Supplement 2), this is probably accurate for the VHA population. But it is difficult to assess the accuracy of this assumption in the U.S. Even so, this assumption seemed more plausible than that of previous studies that the frequency of G202A is constant, regardless of HbA1c measurement (eMethods and eFigure 10 in Supplement 2).^6,8,24^ Fifth, -α3.7 is difficult to impute and its R^2^ from MVP imputation was only 0.773, but that should result in false negatives rather than false positives.^11,25,26^ Finally, due to the paucity in capturing pre-diabetes by ICD codes in the CDW, we could not determine whether G202A carriers had been clinically designated as having pre-diabetes. Consequently, we may have over-estimated the proportion of patients in pre-diabetes who are misguided by HbA1c as having normoglycemia due to G202A. A conservative estimate of such individuals would still be well over a million, based on less than 20% in NHANES having been told they have pre-diabetes.

G6PD deficiency is a condition especially common to African ancestry and largely asymptomatic and undiagnosed. The presence of G202A and other non-glycemic erythropoietic genetic variants could contribute to underestimating the severity of glycemic dysregulation and delayed management. Our estimates for rates of potentially missed pre-diabetes based on HbA1c alone in females were almost as large as in males. After excluding diagnosed G6PD deficiency, 11% of male Veterans with African ancestry had the G202A variant, which means that their HbA1c level was 0.8 percentage points too low to be an accurate reflection of their glycemic dysregulation. The U.S. African-American adult male population is likely to have a similar percentage of G202A carriers without diagnosis of G6PD deficiency. Further study is warranted to investigate whether this contributes to racial and other disparities in glycemic management.

## Data Availability

Access to individual level data used in the present work requires approval by the U.S. Veterans Administration.

## ACKNOWLEDGEMENTS

This research is based on data from the Million Veteran Program, Office of Research and Development, Veterans Health Administration, and was supported by award CSP#2012 LEAP Initiative, BX003362 from VA Office of R&D and Million Veteran Program-MVP003. This publication does not represent the views of the Department of Veteran Affairs or the United States Government. The authors have no potential conflicts of interest to disclose.

## DATA AVAILABILITY

Access to MVP individual level data requires permission of the VA. All results of our statistical analysis are included in this manuscript and its supplements.

