## Supplement 2 for "Non-glycemic genetic effects on HbA1c and clinical glycemic status in African ancestry: VA Million Veteran Program"

### SUPPLEMENTAL MATERIAL

**Contents**

eMethods
- Ranges of random blood glucose used in definition of glycemic status
- Method to estimate misclassified glycemic status due to the G202A variant effect on HbA1c level
- Changes in variant effects during 2012-2021
- References

eFigures
- eFigure 1. Cohort inclusion and exclusion criteria
- eFigure 2. Distributions of random glucose and HbA1c in males
- eFigure 3. Distributions of random glucose and HbA1c in females
- eFigure 4. Distributions of insulin resistance features among males with HbA1c in pre-diabetes range
- eFigure 5. Among males, non-carriers with HbA1c in pre-diabetes range had insulin resistance features similar to G202A carriers with HbA1c 0.9% lower
- eFigure 6. Distributions of insulin resistance features among females with HbA1c in pre-diabetes range
- eFigure 7. Among females, non-carriers with HbA1c in pre-diabetes range had insulin resistance features similar to G202A homozygous carriers with HbA1c 0.7% lower
- eFigure 8. Estimated variant effects on HbA1c in males as a function of year of measurement
- eFigure 9. Distributions of HbA1c in three populations
- eFigure 10. Frequency of G202A variant as function of sex, age and HbA1c

Nonauthor collaborators. Million Veteran Program Core members

#### eMethods

*Ranges of random blood glucose used in definition of glycemic status.* The WHO T2D criterion for random blood glucose is 200 mg/dL or greater. Only 5% of our cohort had glucose ≥200 mg/dL and 8% had glucose 140-199 mg/dL. On the other hand, 19% had glucose > 125 mg/dL and 32% had glucose 100-125 mg/dL. Finally, 25% had HbA1c ≥6.5% and 43% had HbA1c 5.7-6.4%. We tested for differences in rates of elevated HbA1c between carriers and non-carriers (Table 1A), and then compared those results to differences in rates of elevated glucose (Table 1B). We chose to use the criterion for *fasting* blood glucose so that the rates of elevated glucose were closer to the rates of elevated HbA1c. This strengthens our statistical argument. Likewise, the *fasting* blood glucose criteria gave much more informative distributions in the plots of insulin resistance features (Figure 2; eFigures 5-8 in this Supplement).

*Method to estimate misclassified glycemic status due to the G202A variant effect on HbA1c level.* To help justify our underlying assumptions, we confirmed that the distributions of HbA1c levels were similar between our study and the general VHA patients, as well as between our study and the estimated distributions in NHANES (eFigure 10). Furthermore, eFigure 11 shows the percentage of G202A carriers as a function of sex and age in our MVP cohort, which we assumed to hold in the general VHA patient population and among all non-diabetic African American adults.

*Changes in variant effects during 2012-2021.* Previously reported erythropoietic variant effects on HbA1c, particularly reports of effect size -0.3% for HbS, could be due to interference in older, obsolete assays[1-5]. Since our study began in 2011, we might have included HbA1c measurements from assays with interference. To investigate this, we analyzed whether effect sizes changed over time, using one lab panel for each individual in each year 2012-2021, regardless of enrollment date; in each of these years, 15-25% of each variant group had a lab panel. We compared across years the estimated variant effect on HbA1c between male carriers and male non-carriers. For HbS, there was no overlap between the range of yearly effect sizes in 2012-2017 and the range in 2018-2021 (eFigure 8). In linear regression as for Table 2D of individuals with index date in 2018 or later, HbS estimated effect size for males was -0.135% (SE 0.016%, N=891). The VHA CDW does not specify the assay used for a measurement, but these results suggest improvement in HbS assay interference after 2012.[4]

**eFigures**

#### eFigure 1. Cohort inclusion and exclusion criteria

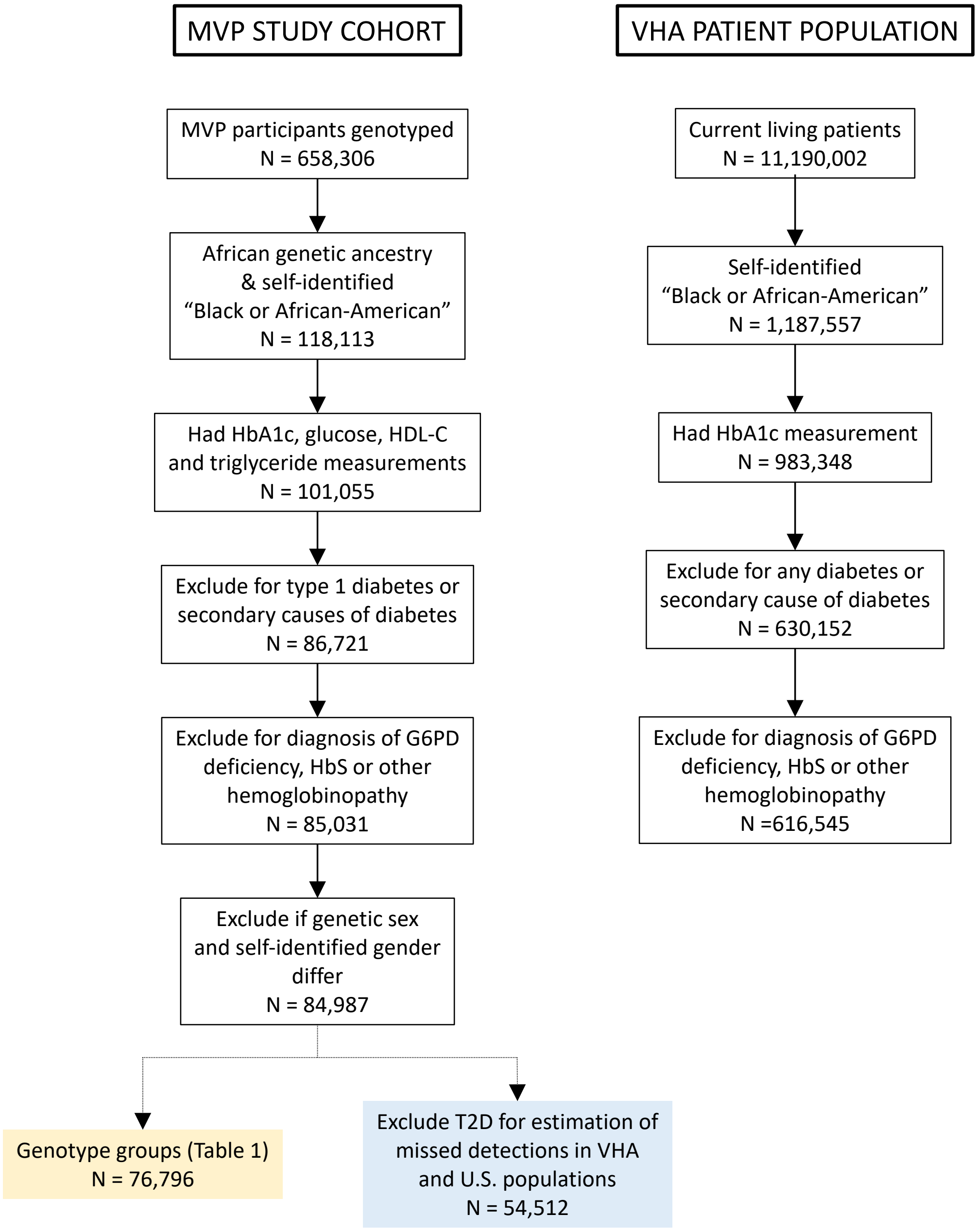

#### eFigure 2. Distributions of random glucose and HbA1c in males

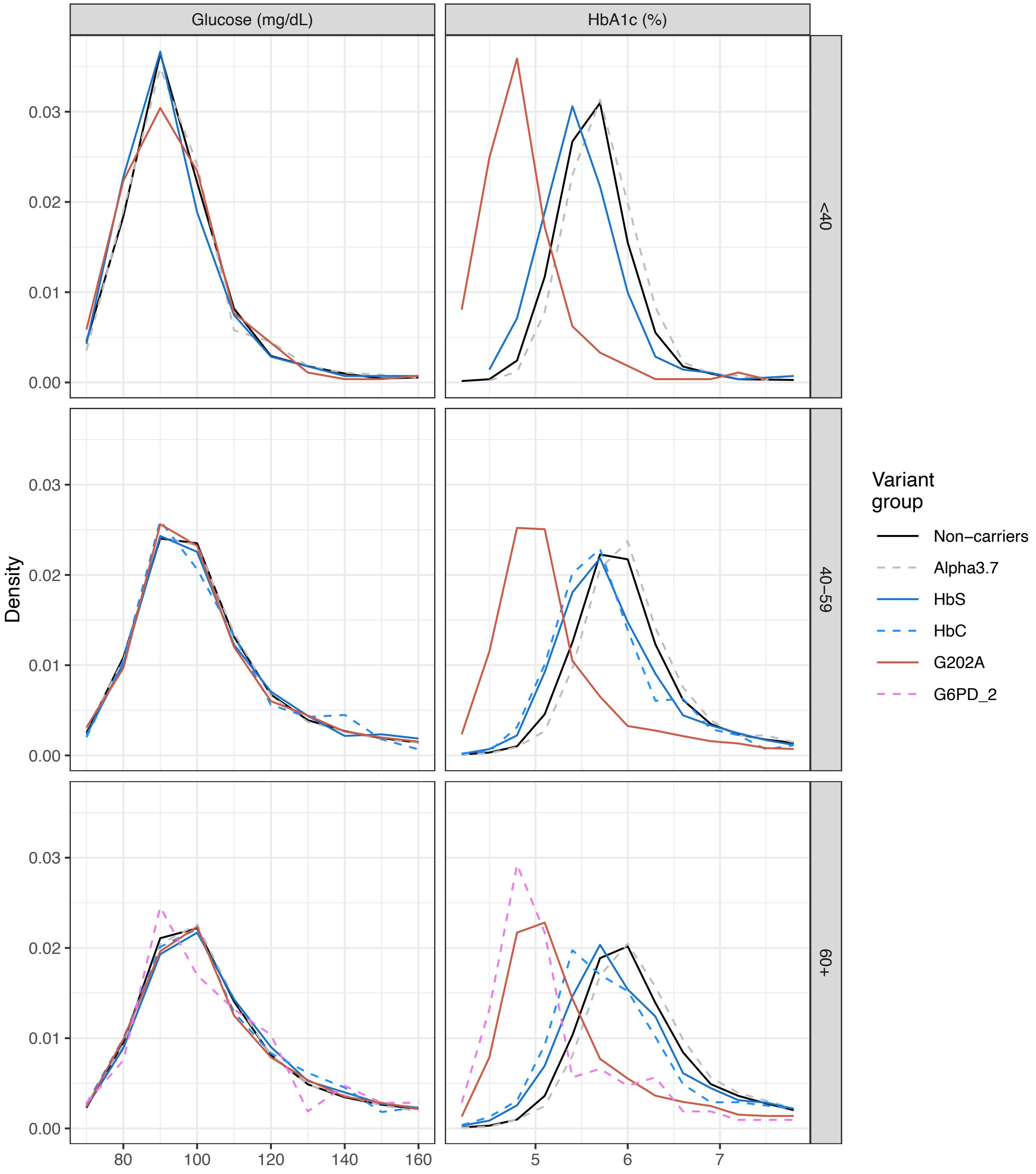


Each panel shows distributions for given age and variant group, omitting distributions based on fewer than 100 individuals.

#### eFigure 3. Distributions of random glucose and HbA1c in females

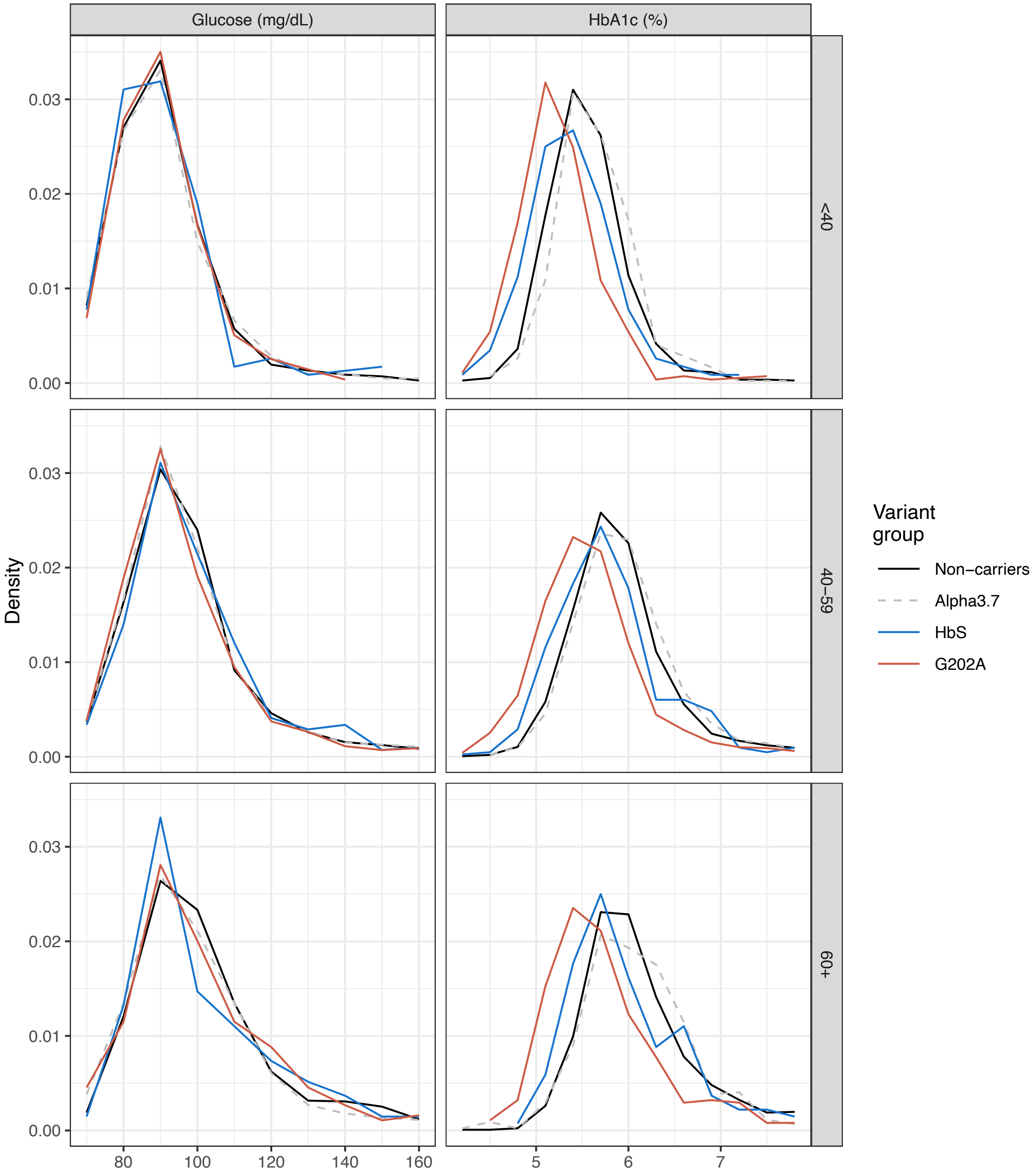


Each panel shows distributions for given age and variant group, omitting distributions based on fewer than 100 individuals.

#### eFigure 4. Distributions of insulin resistance features among males with HbA1c in pre-diabetes range

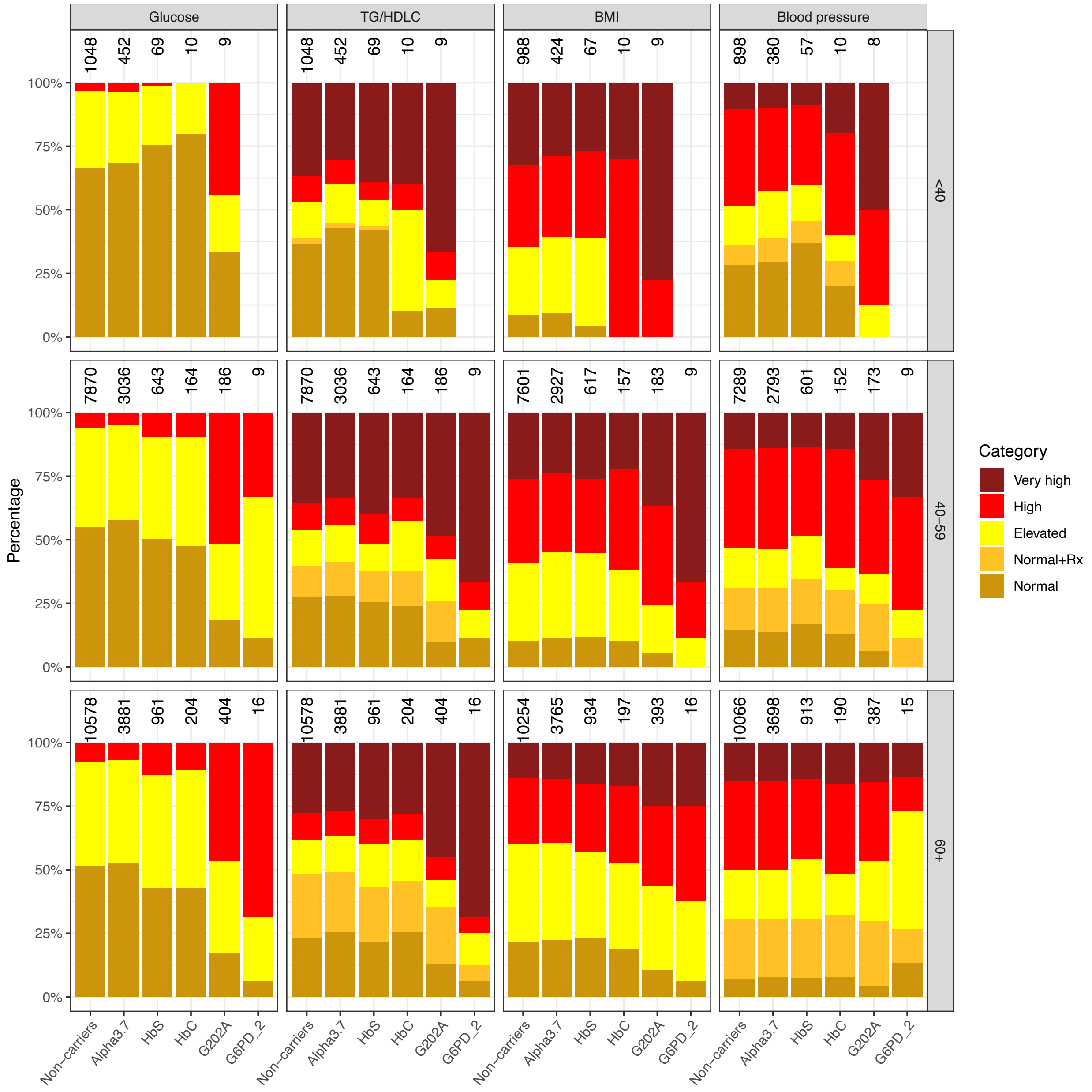


Each panel shows distributions for given age and feature among males with HbA1c in pre-diabetes range (5.7-6.4%). The number above each bar indicates the number of individuals represented, omitting bars for fewer than five individuals. Categories of insulin resistance features defined in eTable 4 in Supplement 2.

#### eFigure 5. Among males, non-carriers with HbA1c in pre-diabetes range had insulin resistance features similar to G202A carriers with HbA1c 0.8% lower

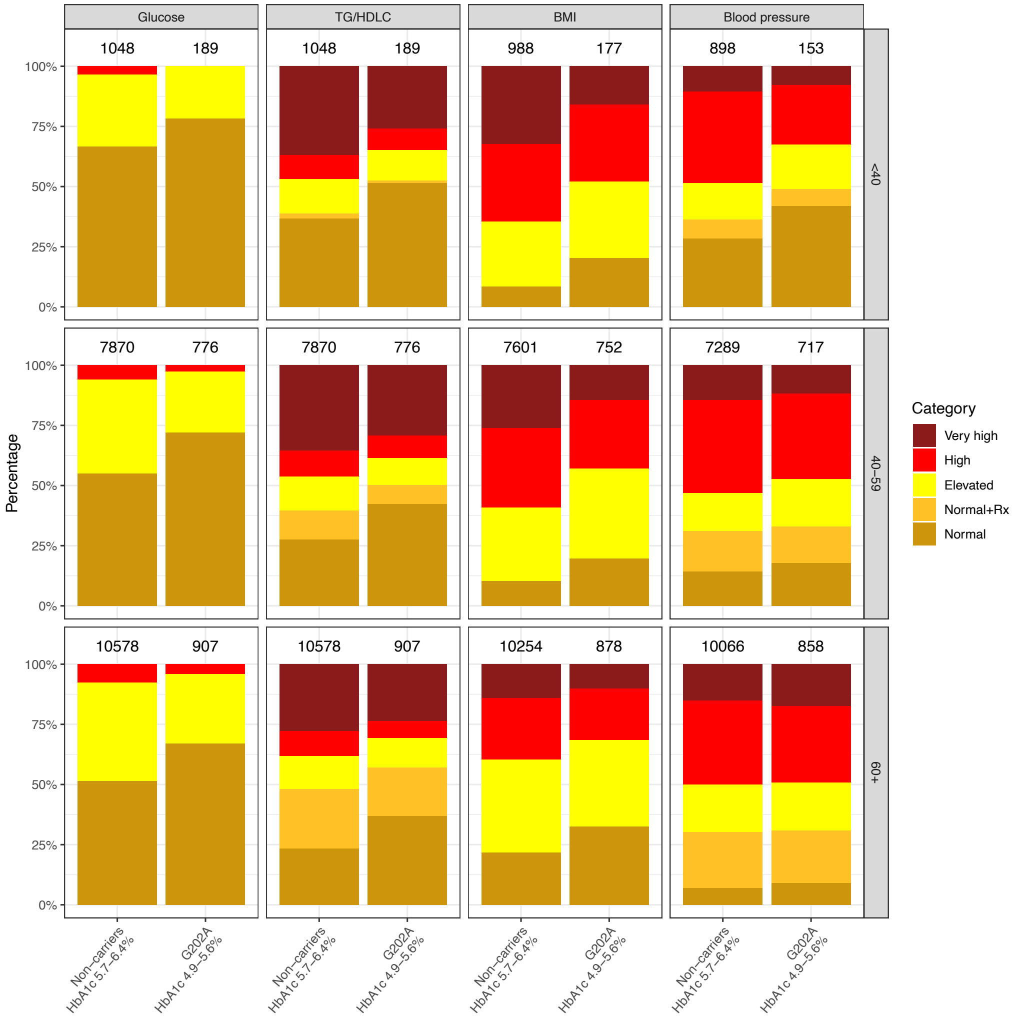


Each panel shows distributions for given age and feature among non-carriers with HbA1c 5.7-6.4% and G202A carriers with HbA1c 4.8-5.5%. The number above each bar indicates the number of individuals represented. Categories of insulin resistance features defined in eTable 4 in Supplement 2.

**eFigure 6. Distributions of insulin resistance features among females with HbA1c in pre-diabetes range**

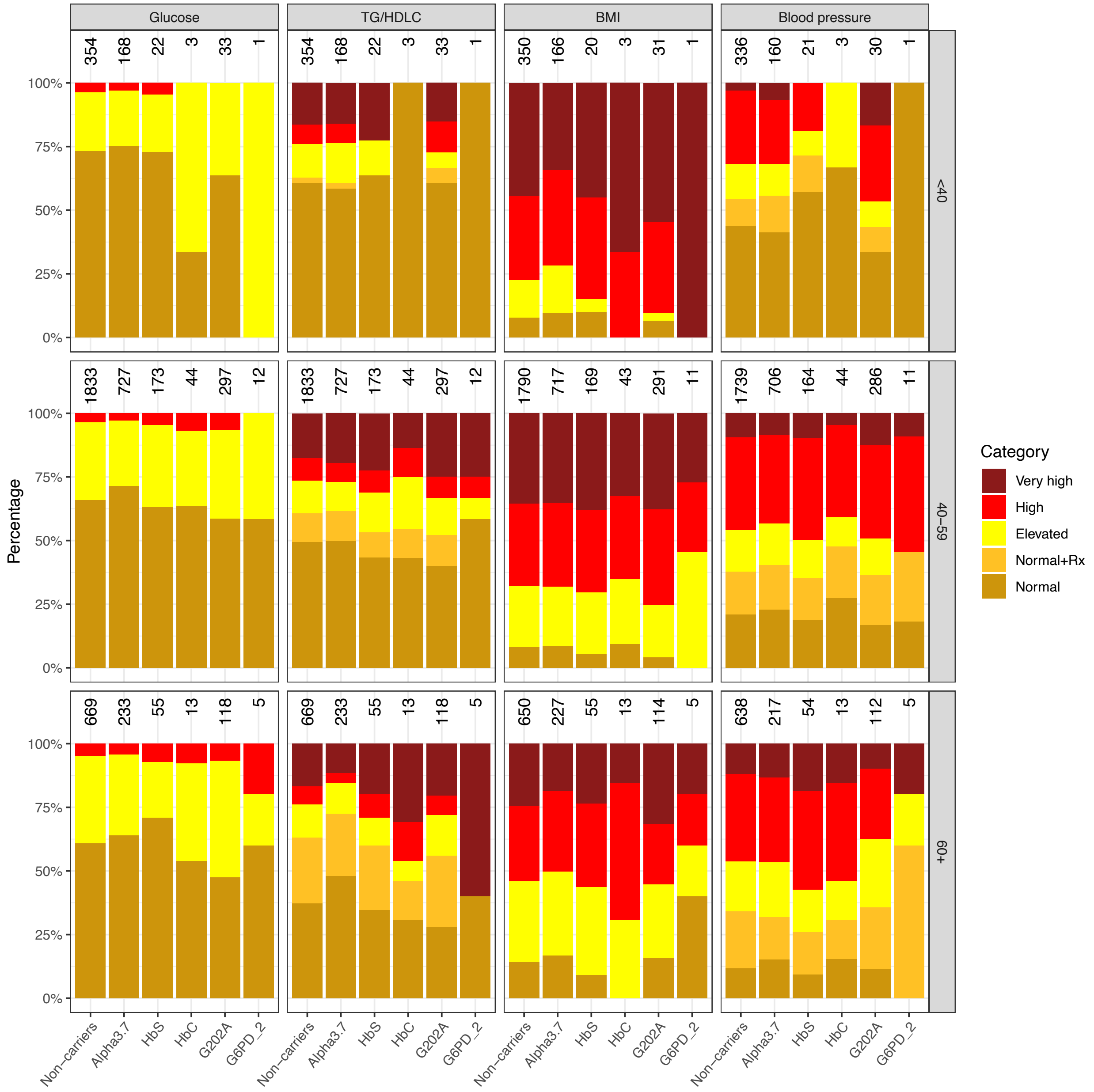


Each panel shows distributions for given age and feature among females with HbA1c in pre-diabetes range (5.7-6.4%). The number above each bar indicates the number of individuals represented. Categories of insulin resistance features defined in eTable 4 in Supplement 2.

#### eFigure 7. Among females, non-carriers with HbA1c in pre-diabetes range had insulin resistance features similar to G202A homozygous carriers with HbA1c 0.7% lower

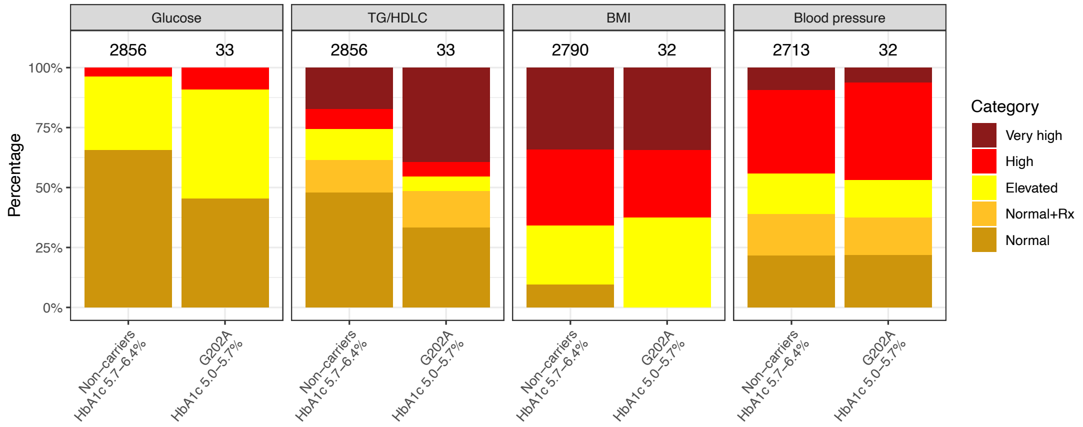


Each panel shows distributions for given feature among females 40 years or older who are either non-carriers with HbA1c 5.7-6.4% or G202A homozygous carriers with HbA1c 5.0-5.7%. The number above each bar indicates the number of individuals represented. Categories of insulin resistance features defined in eTable 4 in Supplement 2.

#### eFigure 8. Estimated variant effects on HbA1c in males as a function of year of measurement

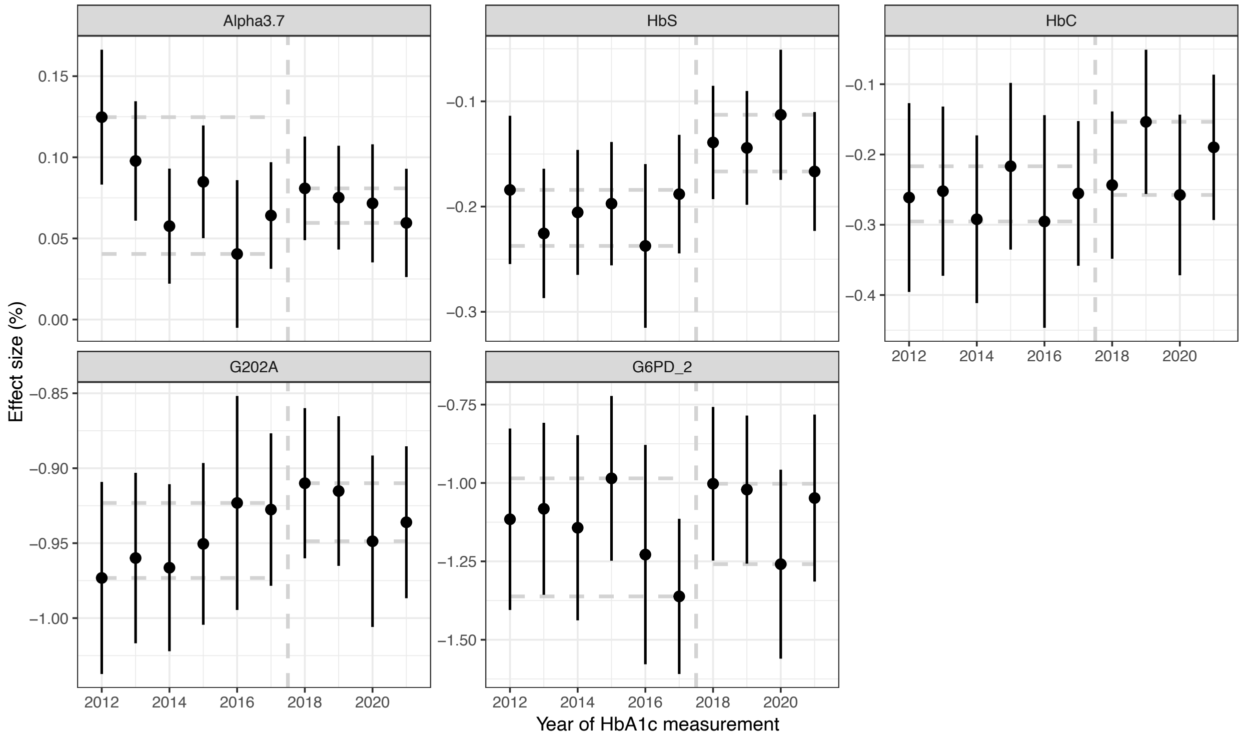


Vertical lines indicate ±2 standard errors. Horizontal dashed lines indicate range of effect sizes in 2012-2017 and in 2018-2021. Vertical dashed line separates the two time periods.

#### eFigure 9. Distributions of HbA1c in three populations

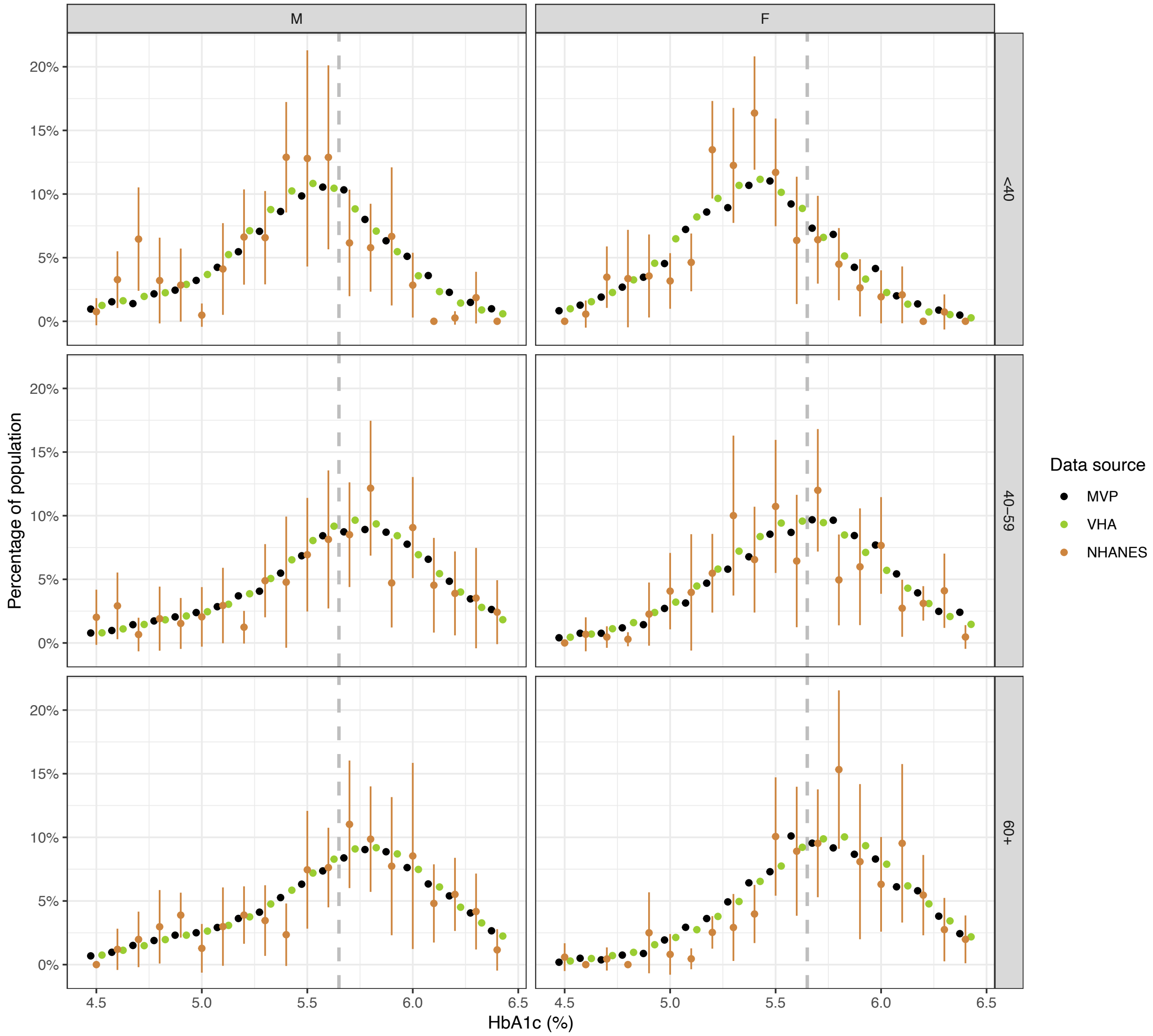


Each panel shows distributions for given age and sex among non-diabetic African American adults in (1) MVP study cohort, (2) general VHA patients, and (3) U.S. at large. The last was based on NHANES survey and shows 95% confidence intervals incorporating sampling error. The dashed vertical line separates normal and pre-diabetes ranges of HbA1c. Among males under 40, the distribution from NHANES had a bump around 4.7%, near the median of 4.8% in the MVP G202A group of same age.

#### eFigure 10. Frequency of G202A variant as function of sex, age and HbA1c

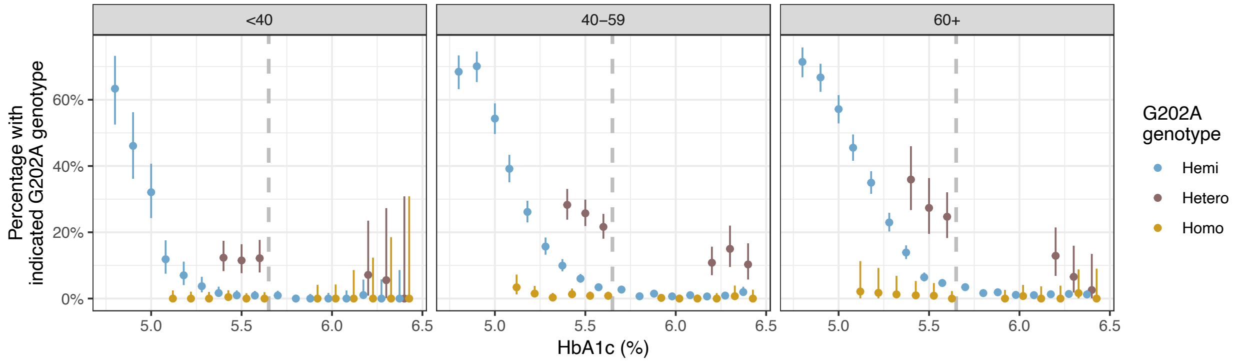


In MVP cohort without diabetes diagnosis or medication, percentage of individuals that had G202A variant as a function of their HbA1c level, grouped by sex, age and number of G202A variants.

**Million Veteran Program Core members**

MVP Program Office

- Program Director - Sumitra Muralidhar, Ph.D.
  US Department of Veterans Affairs, 810 Vermont Avenue NW, Washington, DC 20420
- Associate Director, Scientific Programs - Jennifer Moser, Ph.D.
  US Department of Veterans Affairs, 810 Vermont Avenue NW, Washington, DC 20420
- Associate Director, Cohort Management & Public Relations - Jennifer E. Deen, B.S.
  US Department of Veterans Affairs, 810 Vermont Avenue NW, Washington, DC 20420

MVP Executive Committee

- Co-Chair: J. Michael Gaziano, M.D., M.P.H.
  VA Boston Healthcare System, 150 S. Huntington Avenue, Boston, MA 02130
- Co-Chair: Sumitra Muralidhar, Ph.D.
  US Department of Veterans Affairs, 810 Vermont Avenue NW, Washington, DC 20420
- Jean Beckham, Ph.D.
  Durham VA Medical Center, 508 Fulton Street, Durham, NC 27705
- Kyong-Mi Chang, M.D.
  Philadelphia VA Medical Center, 3900 Woodland Avenue, Philadelphia, PA 19104
- Philip S. Tsao, Ph.D.
  VA Palo Alto Health Care System, 3801 Miranda Avenue, Palo Alto, CA 94304
- Shiuh-Wen Luoh, M.D., Ph.D.
  VA Portland Health Care System, 3710 SW US Veterans Hospital Rd, Portland, OR 97239
  US Department of Veterans Affairs, 810 Vermont Avenue NW, Washington, DC 20420
- Juan P. Casas, M.D., Ph.D., Ex-Officio
  VA Boston Healthcare System, 150 S. Huntington Avenue, Boston, MA 02130

MVP Principal Investigators

- J. Michael Gaziano, M.D., M.P.H.
  VA Boston Healthcare System, 150 S. Huntington Avenue, Boston, MA 02130
- Philip S. Tsao, Ph.D.
  VA Palo Alto Health Care System, 3801 Miranda Avenue, Palo Alto, CA 94304

MVP Operations

- MVP Executive Director – Juan P. Casas, M.D., Ph.D.
  VA Boston Healthcare System, 150 S. Huntington Avenue, Boston, MA 02130
- Director of Regulatory Affairs – Lori Churby, B.S.
  VA Palo Alto Health Care System, 3801 Miranda Avenue, Palo Alto, CA 94304
- MVP Cohort Management Director – Stacey B. Whitbourne, Ph.D.
  VA Boston Healthcare System, 150 S. Huntington Avenue, Boston, MA 02130
- MVP Recruitment/Enrollment Director - Jessica V. Brewer, M.P.H.
  VA Boston Healthcare System, 150 S. Huntington Avenue, Boston, MA 02130
- Director, VA Central Biorepository, Boston – Mary T. Brophy M.D., M.P.H.
  VA Boston Healthcare System, 150 S. Huntington Avenue, Boston, MA 02130
- Executive Director for MVP Biorepositories - Luis E. Selva, Ph.D.
  VA Boston Healthcare System, 150 S. Huntington Avenue, Boston, MA 02130
- MVP Informatics, Boston – Shahpoor (Alex) Shayan, M.S.
  VA Boston Healthcare System, 150 S. Huntington Avenue, Boston, MA 02130
- Director, MVP Data Operations/Analytics, Boston – Kelly Cho, M.P.H., Ph.D.
  VA Boston Healthcare System, 150 S. Huntington Avenue, Boston, MA 02130
- Director, Center for Computational and Data Science (C-DACS) & Genomics Core – Saiju Pyarajan Ph.D.
  VA Boston Healthcare System, 150 S. Huntington Avenue, Boston, MA 02130
- Director, Molecular Data Core – Philip S. Tsao, Ph.D.
  VA Palo Alto Health Care System, 3801 Miranda Avenue, Palo Alto, CA 94304
- Director, Phenomics Data Core – Kelly Cho, M.P.H, Ph.D.

VA Boston Healthcare System, 150 S. Huntington Avenue, Boston, MA 02130
- Director, VA Informatics and Computing Infrastructure (VINCI) – Scott L. DuVall, Ph.D.
  VA Salt Lake City Health Care System, 500 Foothill Drive, Salt Lake City, UT 84148
- MVP Coordinating Centers
  - Cooperative Studies Program Clinical Research Pharmacy Coordinating Center, Albuquerque – Todd Connor, Pharm.D.; Dean P. Argyres, B.S., M.S.
    New Mexico VA Health Care System, 1501 San Pedro Drive SE, Albuquerque, NM 87108
  - Genomics Coordinating Center, Palo Alto – Philip S. Tsao, Ph.D.
    VA Palo Alto Health Care System, 3801 Miranda Avenue, Palo Alto, CA 94304
  - MVP Boston Coordinating Center, Boston - J. Michael Gaziano, M.D., M.P.H.
    VA Boston Healthcare System, 150 S. Huntington Avenue, Boston, MA 02130
  - MVP Information Center, Canandaigua – Brady Stephens, M.S.
    Canandaigua VA Medical Center, 400 Fort Hill Avenue, Canandaigua, NY 14424

Current MVP Local Site Investigators
- Atlanta VA Medical Center (Peter Wilson, M.D.)
  1670 Clairmont Road, Decatur, GA 30033
- Bay Pines VA Healthcare System (Rachel McArdle, Ph.D.)
  10,000 Bay Pines Blvd Bay Pines, FL 33744
- Birmingham VA Medical Center (Louis Dellitalia, M.D.)
  700 S. 19th Street, Birmingham AL 35233
- Central Western Massachusetts Healthcare System (Kristin Mattocks, Ph.D., M.P.H.)
  421 North Main Street, Leeds, MA 01053
- Cincinnati VA Medical Center (John Harley, M.D., Ph.D.)
  3200 Vine Street, Cincinnati, OH 45220
- Clement J. Zablocki VA Medical Center (Jeffrey Whittle, M.D., M.P.H.)
  5000 West National Avenue, Milwaukee, WI 53295
- VA Northeast Ohio Healthcare System (Frank Jacono, M.D.)
  10701 East Boulevard, Cleveland, OH 44106
- Durham VA Medical Center (Jean Beckham, Ph.D.)
  508 Fulton Street, Durham, NC 27705
- Edith Nourse Rogers Memorial Veterans Hospital (John Wells., Ph.D.)
  200 Springs Road, Bedford, MA 01730
- Edward Hines, Jr. VA Medical Center (Salvador Gutierrez, M.D.)
  5000 South 5th Avenue, Hines, IL 60141
- Veterans Health Care System of the Ozarks (Kathrina Alexander, M.D.)
  1100 North College Avenue, Fayetteville, AR 72703
- Fargo VA Health Care System (Kimberly Hammer, Ph.D.)
  2101 N. Elm, Fargo, ND 58102
- VA Health Care Upstate New York (James Norton, Ph.D.)
  113 Holland Avenue, Albany, NY 12208
- New Mexico VA Health Care System (Gerardo Villareal, M.D.)
  1501 San Pedro Drive, S.E. Albuquerque, NM 87108
- VA Boston Healthcare System (Scott Kinlay, M.B.B.S., Ph.D.)
  150 S. Huntington Avenue, Boston, MA 02130
- VA Western New York Healthcare System (Junzhe Xu, M.D.)
  3495 Bailey Avenue, Buffalo, NY 14215-1199
- Ralph H. Johnson VA Medical Center (Mark Hamner, M.D.)
  109 Bee Street, Mental Health Research, Charleston, SC 29401
- Columbia VA Health Care System (Roy Mathew, M.D.)
  6439 Garners Ferry Road, Columbia, SC 29209
- VA North Texas Health Care System (Sujata Bhushan, M.D.)
  4500 S. Lancaster Road, Dallas, TX 75216

- Hampton VA Medical Center (Pran Iruvanti, D.O., Ph.D.)
  100 Emancipation Drive, Hampton, VA 23667
- Richmond VA Medical Center (Michael Godschalk, M.D.)
  1201 Broad Rock Blvd., Richmond, VA 23249
- Iowa City VA Health Care System (Zuhair Ballas, M.D.)
  601 Highway 6 West, Iowa City, IA 52246-2208
- Eastern Oklahoma VA Health Care System (River Smith, Ph.D.)
  1011 Honor Heights Drive, Muskogee, OK 74401
- James A. Haley Veterans' Hospital (Stephen Mastorides, M.D.)
  13000 Bruce B. Downs Blvd, Tampa, FL 33612
- James H. Quillen VA Medical Center (Jonathan Moorman, M.D., Ph.D.)
  Corner of Lamont & Veterans Way, Mountain Home, TN 37684
- John D. Dingell VA Medical Center (Saib Gappy, M.D.)
  4646 John R Street, Detroit, MI 48201
- Louisville VA Medical Center (Jon Klein, M.D., Ph.D.)
  800 Zorn Avenue, Louisville, KY 40206
- Manchester VA Medical Center (Nora Ratcliffe, M.D.)
  718 Smyth Road, Manchester, NH 03104
- Miami VA Health Care System (Ana Palacio, M.D., M.P.H.)
  1201 NW 16th Street, 11 GRC, Miami FL 33125
- Michael E. DeBakey VA Medical Center (Olaoluwa Okusaga, M.D.)
  2002 Holcombe Blvd, Houston, TX 77030
- Minneapolis VA Health Care System (Maureen Murdoch, M.D., M.P.H.)
  One Veterans Drive, Minneapolis, MN 55417
- N. FL/S. GA Veterans Health System (Peruvemba Sriram, M.D.)
  1601 SW Archer Road, Gainesville, FL 32608
- Northport VA Medical Center (Shing Shing Yeh, Ph.D., M.D.)
  79 Middleville Road, Northport, NY 11768
- Overton Brooks VA Medical Center (Neeraj Tandon, M.D.)
  510 East Stoner Ave, Shreveport, LA 71101
- Philadelphia VA Medical Center (Darshana Jhala, M.D.)
  3900 Woodland Avenue, Philadelphia, PA 19104
- Phoenix VA Health Care System (Samuel Aguayo, M.D.)
  650 E. Indian School Road, Phoenix, AZ 85012
- Portland VA Medical Center (David Cohen, M.D.)
  3710 SW U.S. Veterans Hospital Road, Portland, OR 97239
- Providence VA Medical Center (Satish Sharma, M.D.)
  830 Chalkstone Avenue, Providence, RI 02908
- Richard Roudebush VA Medical Center (Suthat Liangpunsakul, M.D., M.P.H.)
  1481 West 10th Street, Indianapolis, IN 46202
- Salem VA Medical Center (Kris Ann Oursler, M.D.)
  1970 Roanoke Blvd, Salem, VA 24153
- San Francisco VA Health Care System (Mary Whooley, M.D.)
  4150 Clement Street, San Francisco, CA 94121
- South Texas Veterans Health Care System (Sunil Ahuja, M.D.)
  7400 Merton Minter Boulevard, San Antonio, TX 78229
- Southeast Louisiana Veterans Health Care System (Joseph Constans, Ph.D.)
  2400 Canal Street, New Orleans, LA 70119
- Southern Arizona VA Health Care System (Paul Meyer, M.D., Ph.D.)
  3601 S 6th Avenue, Tucson, AZ 85723
- Sioux Falls VA Health Care System (Jennifer Greco, M.D.)
  2501 W 22nd Street, Sioux Falls, SD 57105

- St. Louis VA Health Care System (Michael Rauchman, M.D.)
  915 North Grand Blvd, St. Louis, MO 63106
- Syracuse VA Medical Center (Richard Servatius, Ph.D.)
  800 Irving Avenue, Syracuse, NY 13210
- VA Eastern Kansas Health Care System (Melinda Gaddy, Ph.D.)
  4101 S 4th Street Trafficway, Leavenworth, KS 66048
- VA Greater Los Angeles Health Care System (Agnes Wallbom, M.D., M.S.)
  11301 Wilshire Blvd, Los Angeles, CA 90073
- VA Long Beach Healthcare System (Timothy Morgan, M.D.)
  5901 East 7th Street Long Beach, CA 90822
- VA Maine Healthcare System (Todd Stapley, D.O.)
  1 VA Center, Augusta, ME 04330
- VA New York Harbor Healthcare System (Peter Liang, M.D., M.P.H.)
  423 East 23rd Street, New York, NY 10010
- VA Pacific Islands Health Care System (Daryl Fujii, Ph.D.)
  459 Patterson Rd, Honolulu, HI 96819
- VA Palo Alto Health Care System (Philip Tsao, Ph.D.)
  3801 Miranda Avenue, Palo Alto, CA 94304-1290
- VA Pittsburgh Health Care System (Patrick Strollo, Jr., M.D.)
  University Drive, Pittsburgh, PA 15240
- VA Puget Sound Health Care System (Edward Boyko, M.D.)
  1660 S. Columbian Way, Seattle, WA 98108-1597
- VA Salt Lake City Health Care System (Jessica Walsh, M.D.)
  500 Foothill Drive, Salt Lake City, UT 84148
- VA San Diego Healthcare System (Samir Gupta, M.D., M.S.C.S.)
  3350 La Jolla Village Drive, San Diego, CA 92161
- VA Sierra Nevada Health Care System (Mostaqul Huq, Pharm.D., Ph.D.)
  975 Kirman Avenue, Reno, NV 89502
- VA Southern Nevada Healthcare System (Joseph Fayad, M.D.)
  6900 North Pecos Road, North Las Vegas, NV 89086
- VA Tennessee Valley Healthcare System (Adriana Hung, M.D., M.P.H.)
  1310 24th Avenue, South Nashville, TN 37212
- Washington DC VA Medical Center (Jack Lichy, M.D., Ph.D.)
  50 Irving St, Washington, D. C. 20422
- W.G. (Bill) Hefner VA Medical Center (Robin Hurley, M.D.)
  1601 Brenner Ave, Salisbury, NC 28144
- White River Junction VA Medical Center (Brooks Robey, M.D.)
  163 Veterans Drive, White River Junction, VT 05009
- William S. Middleton Memorial Veterans Hospital (Prakash Balasubramanian, M.D.)
  2500 Overlook Terrace, Madison, WI 53705
